# Development and Validation of a Convolutional Neural Network Model for ICU Acute Kidney Injury Prediction

**DOI:** 10.1101/2020.05.01.20087551

**Authors:** Sidney Le, Angier Allen, Jacob Calvert, Paul M. Palevsky, Gregory Braden, Sharad Patel, Emily Pellegrini, Abigail Green-Saxena, Jana Hoffman, Ritankar Das

## Abstract

**Rationale and objectives:** Acute kidney injury (AKI) is common among hospitalized patients and has a significant impact on morbidity and mortality. While early prediction of AKI has the potential to reduce adverse patient outcomes, it remains a difficult condition to predict and diagnose. The purpose of this study was to evaluate the ability of a machine learning algorithm to predict for AKI KDIGO Stage 2 or 3 up to 72 hours in advance of onset using convolutional recurrent neural nets (CNN) and patient Electronic Health Record (EHR) data.

**Methods:** A CNN prediction system was developed to continuously and automatically monitor for incipient AKI. 7122 patient encounters were retrospectively analyzed from the Medical Information Mart for Intensive Care III (MIMIC-III) database.

**New Predictors and Established Predictors:** New predictor - CNN machine learning-based AKI prediction model. Established predictors - XGBoost AKI prediction model and the Sequential Organ Failure Assessment (SOFA) scoring system.

**Outcomes:** AKI onset.

**Analytical Approach:** The model was trained on routinely-collected patient EHR data. Measurements included Area Under the Receiver Operating Characteristic (AUROC) curve, positive predictive value (PPV), and a battery of additional performance metrics for 72 hour advance prediction of AKI onset.

**Results:** On a hold-out test set, the algorithm attained an AUROC of 0.85 and PPV of 0.25, relative to a cohort AKI prevalence of 5.21%, for long-horizon AKI prediction at a 72-hour window prior to onset.

**Conclusions:** A CNN machine learning-based AKI prediction model outperforms XGBoost and the SOFA scoring system, demonstrating superior performance in predicting acute kidney injury 72 hours prior to onset, without reliance on changes in serum creatinine.

## INTRODUCTION

Acute kidney injury (AKI) is a complex syndrome associated with large clinical and financial burdens [1–12]. Despite its prevalence in hospitalized patients [2,13] and reported incidence as high as 70% in the critically ill [13,14], no treatment has been developed to effectively reverse injury to the kidney and restore kidney function [1]. The reasons for this failure have been attributed to delays in diagnosis and intervention [2, 15–23], the complex nature of the AKI syndrome and the staging of its severity [3, 21], and its multiple etiologies [15,16].

Until recently, studies of incidence and outcomes of AKI have produced inconsistent results due to varying definitions of AKI [24–26]. The Risk, Injury, Failure, Loss, End-stage kidney disease (RIFLE) criteria [27], followed by the Acute Kidney Injury Network (AKIN) [28] and most recently the Kidney Disease: Improving Global Outcomes (KDIGO) criteria [29, 30] have provided consensus on an AKI definition. KDIGO guidelines define acute kidney injury as an absolute increase of serum creatinine (SCr) of >0·3 mg/dL within 48 hours or a relative increase of >50% over no more than 7 days [21, 29]. Doubling of SCr at steady state reflects an approximate 50% decrease in kidney function as assessed by glomerular filtration rate (GFR) [31]. Some studies have suggested that changes in SCr even smaller than 0.3 mg/dL within 48 hours are associated with significant increases in the risk of death, dialysis, and other morbidities [6, 21, 32–38], and other studies are consistent with worsening outcomes with increasing AKI stage [5, 24, 39–43]. However, increases of serum creatinine are known to lag kidney injury by hours to days after the initial kidney insult, and therefore recognition of AKI is delayed by reliance on SCr measurements [44,45].

Early AKI detection is critical to improving patient outcomes [46–49]. Given that the components necessary for defining and staging AKI are routinely available in the electronic health record (EHR) [3], a number of automated alerts have been developed to predict AKI events prior to onset. However, these alerts are generally triggered by detecting changes in SCr and/or urine output [17]. Because a range of kidney injury can exist before the loss of kidney function can be estimated with these standard laboratory tests [45,50], there is great interest in developing methods that could be used to detect AKI in patients at an earlier stage [51–57]. In this paper, we describe our methodology for the development of a convolutional neural net prediction system that continuously and automatically monitors for incipient AKI, using patient data extracted from the EHR, without requiring serum creatinine or urine output values.

## MATERIALS AND METHODS

### Description of data

This study uses data from the Multiparameter Intelligent Monitoring in Intensive Care (MIMIC)-III version 1.3 dataset [58], collected at Beth Israel Deaconess Medical Center in Boston, MA from 2001 to 2012. The MIMIC dataset offers a variety of encounter information from more than 40,000 unique patients and includes both structured (e.g. lab results) and unstructured (e.g. clinician notes) data. Due to differences in the storage of patient procedures information, we restrict our study to data collected from 2008 to 2012 using the MetaVision (iMDSoft) EHR system, and do not include data collected from 2001 to 2008 using the CareVue (Philips) system [59]. Because the collection of the MIMIC data did not affect patient safety and because all data were anonymized in accordance with the Health Insurance Portability and Accountability Act (HIPAA) Privacy Rule, the Institutional Review Boards of Beth Israel Deaconess Medical Center and the Massachusetts Institute of Technology have waived the requirement for patient consent.

From the MetaVision EHR MIMIC encounters, we selected for inclusion those stays involving adult patients (i.e. age 18 years or older) with at least one measurement of diastolic blood pressure, systolic blood pressure, temperature, respiratory rate, heart rate, SpO_2_, and Glasgow Coma Scale. These measurements were selected because they are frequently available and easily collected at the patient bedside, even before clinical suspicion of AKI is present. These were the only direct variables used during training and testing of the algorithm. Serum creatinine was used to determine the gold standard of AKI true positive patients, but was not used as an input in testing. To facilitate the analysis of 72-hour advance prediction of AKI onset with a five-hour window of measurements upon which to base such a prediction, we required patient stay duration to be at least 77 hours in length. For convenience and with minimal restriction, we required that patient encounters lasted no more than 1000 hours. Inclusion criteria are listed in **Figure 1** for 24, 28, and 72 hour prediction windows, and the demographic characteristics of encounters meeting the inclusion criteria are reported in **Table 1**.

**Figure 1.**
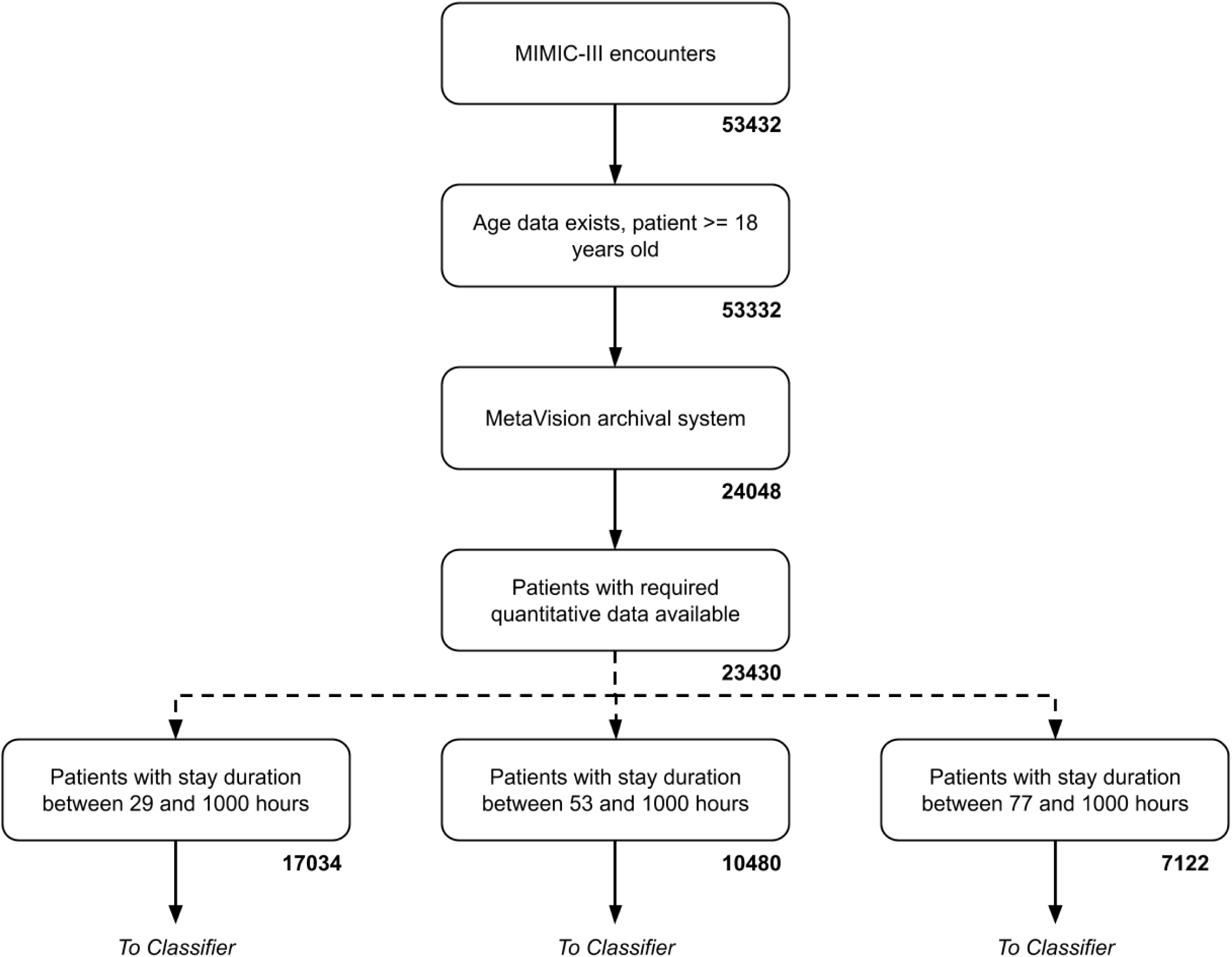
Inclusion diagram.

**Table 1.** Demographic characteristics of MIMIC III ICU encounters meeting the inclusion criteria of Figure 1. We note that the determination of KDIGO positive or negative was made after the data preprocessing steps described in the Methods section.

| Characteristic |  | Count | Percent |
| --- | --- | --- | --- |
| Gender | Female | 3,126 | 43.89 |
|  | Male | 3,996 | 56.11 |
| Age (days):<br>Median 66, IQR (54-78) | 18-29 | 247 | 1.74 |
|  | 30-39 | 4,081 | 28.73 |
|  | 40-49 | 3,945 | 27.77 |
|  | 50-59 | 1,251 | 8.81 |
|  | 60-69 | 1,694 | 11.93 |
|  | 70+ | 2,987 | 21.03 |
| Length of Stay (days):<br>Median 5,<br>IQR (4-9) | 3-5 | 3,739 | 52.50 |
|  | 6-8 | 1,420 | 19.94 |
|  | 9-11 | 693 | 9.73 |
|  | 12+ | 1,270 | 17.83 |
| In-Hospital Death | Yes | 2,481 | 34.84 |
|  | No | 4,641 | 65.16 |
| KDIGO 2/3 | Positive | 371 | 5.21 |
|  | Negative | 6,751 | 94.79 |
| KDIGO 1/2/3 | Positive | 510 | 7.16 |
|  | Negative | 6,612 | 92.84 |

### Overview of preprocessing, training, and testing

Patient encounters satisfying the inclusion criteria were immediately allocated to training and testing sets. Roughly 90% and 10% of all encounters were randomly allocated to the training (n = 6410 patient encounters) and testing sets (n = 710 patient encounters), respectively, stratifying by positive and negative class to ensure equal representation of classes in both sets. We binned the data by the hour, imputed missing measurements, and standardized measurements on a variable-by-variable basis. KDIGO Stage 2 or Stage 3 classifications were determined for each encounter, along with the corresponding times of KDIGO “onset” where appropriate. Stage 2 AKI is defined in the KDIGO staging system as an increase in SCr to more than 200% to 300% (>2- to 3-fold) from baseline or urine output <0.5 ml/kg per hour for more than 12 hours [29]. Stage 3 AKI is defined as an increase in SCr to more than 300% (>3-fold) from baseline, or ≥ 4.0 mg/dl (≥ 354 mmol/l), or kidney replacement therapy (KRT), or a decrease in estimated glomerular filtration rate (eGFR) to < 35 ml/min per 1.73m^2^ (if <18 years of age), or urine output < 0.5 mL/kg/hr for ≥ 24 hours or anuria for ≥ 12 hours [29].

A Doc2Vec embedding network was created to vectorize clinical text data. The embedding network was prepared on a large collection of mid-stay clinical notes, ranging from primary complaint to radiology notes, including everything up to, but not including, the discharge summary, from encounters allocated to the training set. The network embedded texts into 250-dimensional numeric vectors, which served as inputs to the classifiers, alongside the structured data associated with the stays. Training data were passed to a convolutional neural network (CNN) structure, with hyperparameters optimized on the training set using the Python-based optimization package Talos. After the end of training on each fold, network performance was evaluated using the hold-out test set. Results were reported as the average test set performance across cross-validation folds.

### Structured data preprocessing

Structured data were binned by the hour, with multiple intra-hour measurements of the same variable replaced by their average. Missing measurements were handled separately for training and testing sets using last observation carried forward imputation.

### Document vector encoding network and unstructured data preprocessing

To facilitate the use of unstructured text data alongside the structured inputs, we trained a Doc2Vec [60] embedding network with 250 nodes, trained on 238,468 mid-stay clinical notes. Document vectors were produced for the text data available from each encounter, using 125 epochs -- to better ensure the stability of inferred document vector -- and an initial learning rate of 0.01.

### Training of neural network classifier

We constructed a classifier using the Python deep learning library, Keras, that uses variants of multi-channel, multi-headed attention together with convolutions to extract information from the quantitative time series data. A separate network for handling the document vector produced by the Doc2Vec network was combined downstream through concatenation in a fully-connected output layer. Model hyperparameters were optimized using the Nadam optimizer [61] as implemented in the Keras library with learning rate of 0.0009 and binary cross-entropy loss. A diagram of this neural network architecture is available as **Supplementary Figure 1**.

To fit the weights of the network with 10-fold cross-validation, we split the training data into 10 subsets of roughly equal size, and iteratively used 9 subsets for intra-fold training and the final subset for intra-fold testing.

Model parameters were fit over the course of 50 epochs on the 9 intra-fold training subsets, with evaluation on the final subset. For each iterate, we obtained a receiver operating characteristic (ROC) curve, as well as a battery of performance metrics. We then randomly reset the model parameters before performing another iterate. From cross-validation, we obtained an average ROC curve and average performance metrics, along with standard deviation for the performance metrics. Lastly, we evaluated the performance of the trained network on the original, 10% hold-out test set. These results are presented in comparison with an XGBoost [62] classifier and the Sequential Organ Failure Assessment (SOFA) score [63]. Although the SOFA score was not developed for the purpose of long-horizon AKI prediction, it has previously been shown to independently predict AKI risk and outcomes [64,65], and therefore serves as a validated comparison measure for AKI prediction. The XGBoost classifier was trained on the same training sets -- 5-hour windows of quantitative, clinical EHR data -- and evaluated on the same testing set. XGBoost hyperparameters were tuned using a cross-validated grid search on the training data.

## RESULTS

The demographic characteristics associated with MIMIC III ICU encounters meeting the inclusion criteria of **Figure 1** are provided in **Table 1**. The study population was 56.11% male, with few (1.74%) patients younger than 30 years of age and a substantial percentage of patients aged 70 years or more (21.03%). More than half (52.50%) of patients had stays lasting between 3 and 5 days, with a substantial percentage of patients experiencing stays of 12 days or longer (17.83%). The overall mortality rate was 34.84%, with 5.21% of encounters meeting the criteria for KDIGO Stage 2 or Stage 3 at some point, and 7.16% of stays meeting some stage of the KDIGO criteria at any point during the stay.

The results from 10-fold cross-validation on the 90% training set are reported in **Table 2**. The CNN model with the use of the Doc2Vec embeddings of encounter text data outperformed the XGBoost comparator model and the SOFA score for 72-hour advance prediction of KDIGO Stage 2 or Stage 3 onset. We note that, in order to provide non-summative performance metrics (i.e., the metrics other than area under the receiver operating characteristic (AUROC) curve), we selected an operating point for each model or score which provided a sensitivity nearest 0.80. The CNN model performed better (AUROC of 0.85) when text data were made available through Doc2Vec than when these data were unavailable (AUROC of 0.75). In addition, the quality of prediction was higher for KDIGO Stage 2 or Stage 3 onset, as compared with the prediction of onset for any of KDIGO Stages 1-3 (AUROC of 0.81). For corresponding CNN and XGBoost results without oversampling of the minority class, see **Supplementary Table 1**. The results from 10-fold cross-validation for prediction 48-hours (CNN AUROC of 0.835; PPV 0.236) and 24-hours (CNN AUROC of 0.856; PPV 0.221) prior to onset are reported in **Supplementary Table 2** and **Supplementary Table 3**. Permutation feature importance methods were implemented to provide information on the relative importance of each input variable. Feature importance data are presented in **Supplementary Table 4** and **Supplementary Figure 2**.

**Table 2.** Results from 10-fold cross-validation on the MIMIC III data set. The convolutional neural network (CNN) model is compared with an XGBoost classifier, and the Sequential Organ Failure Assessment (SOFA) score. Additional comparison is made to the CNN model without the use of the Doc2Vec network (i.e., without unstructured text data) and for the prediction of KDIGO criteria of any stage. Abbreviations: area under the receiver operating characteristic (AUROC) curve; diagnostic odds ratio (DOR); positive and negative likelihood ratios (LR+ and LR-, respectively); positive and negative predictive value (PPV and NPV, respectively); standard deviation (SD)

|  | CNN | XGBoost | SOFA | No Doc2Vec | Stage 1 Included | Stage 3 Only |
| --- | --- | --- | --- | --- | --- | --- |
| **AUROC mean (SD)** | 0.855 (0.007) | 0.691 (0.007) | 0.652 | 0.747 (0.006) | 0.815 (0.002) | 0.812 (0.006) |
| **Sensitivity mean (SD)** | 0.802 (0.003) | 0.751 (0.091) | 0.781 | 0.802 (0.003) | 0.800 (0.002) | 0.778 (0.000) |
| **Specificity mean (SD)** | 0.743 (0.019) | 0.486 (0.127) | 0.422 | 0.545 (0.017) | 0.675 (0.015) | 0.770 (0.017) |
| **PPV mean (SD)** | 0.245 (0.014) | 0.130 (0.020) | 0.108 | 0.155 (0.005) | 0.303 (0.010) | 0.112 (0.007) |
| **NPV mean (SD)** | 0.972 (0.001) | 0.954 (0.003) | 0.954 | 0.962 (0.001) | 0.949 (0.001) | 0.986 (0.000) |
| **Accuracy mean (SD)** | 0.748 (0.018) | 0.611 (0.083) | 0.499 | 0.569 (0.016) | 0.693 (0.013) | 0.768 (0.016) |
| **DOR mean (SD)** | 11.827 (1.114) | 3.065 (0.310) | 2.596 | 4.861 (0.383) | 8.345 (0.557) | 11.798 (1.148) |
| **LR+ mean (SD)** | 3.140 (0.232) | 1.507 (0.208) | 1.350 | 1.765 (0.069) | 2.469 (0.113) | 3.400 (0.255) |
| **LR- mean (SD)** | 0.266 (0.006) | 0.495 (0.074) | 0.520 | 0.364 (0.015) | 0.296 (0.007) | 0.289 (0.006) |
| **F1 mean (SD)** | 0.375 (0.016) | 0.215 (0.020) | 0.187 | 0.259 (0.007) | 0.439 (0.010) | 0.194 (0.011) |

The CNN model averaged a positive predictive value (PPV) of 0.25 over cross-validation folds for the 72-hour prediction of KDIGO Stages 2 and 3, compared to average PPVs of 0.13 and 0.11 for XGBoost and the SOFA score, respectively (**Table 2**). The advantage of the CNN mostly vanished (PPV of 0.16) in the absence of text data through Doc2Vec input. The average PPV was highest when the CNN classifier was given access to Doc2Vec input and tasked with 72-hour prediction of KDIGO Stages 1-3 (PPV of 0.30). Relative to the 5.21% prevalence of KDIGO Stages 2 and 3, positive predictions made by the CNN model enriched for KDIGO Stage 2 or 3 encounters by a factor of 4.80, whereas XGBoost and the SOFA score enriched these encounters by factors of 2.50 and 2.11, respectively.

The ROC curve comparison of 72-hour prediction on the 10% hold-out test set is shown in **Figure 2**. The CNN model, which was provided text data through Doc2Vec input, performed substantially better than the XGBoost model and the SOFA score. The XGBoost model and SOFA had similar performance on the test set.

**Figure 2.**
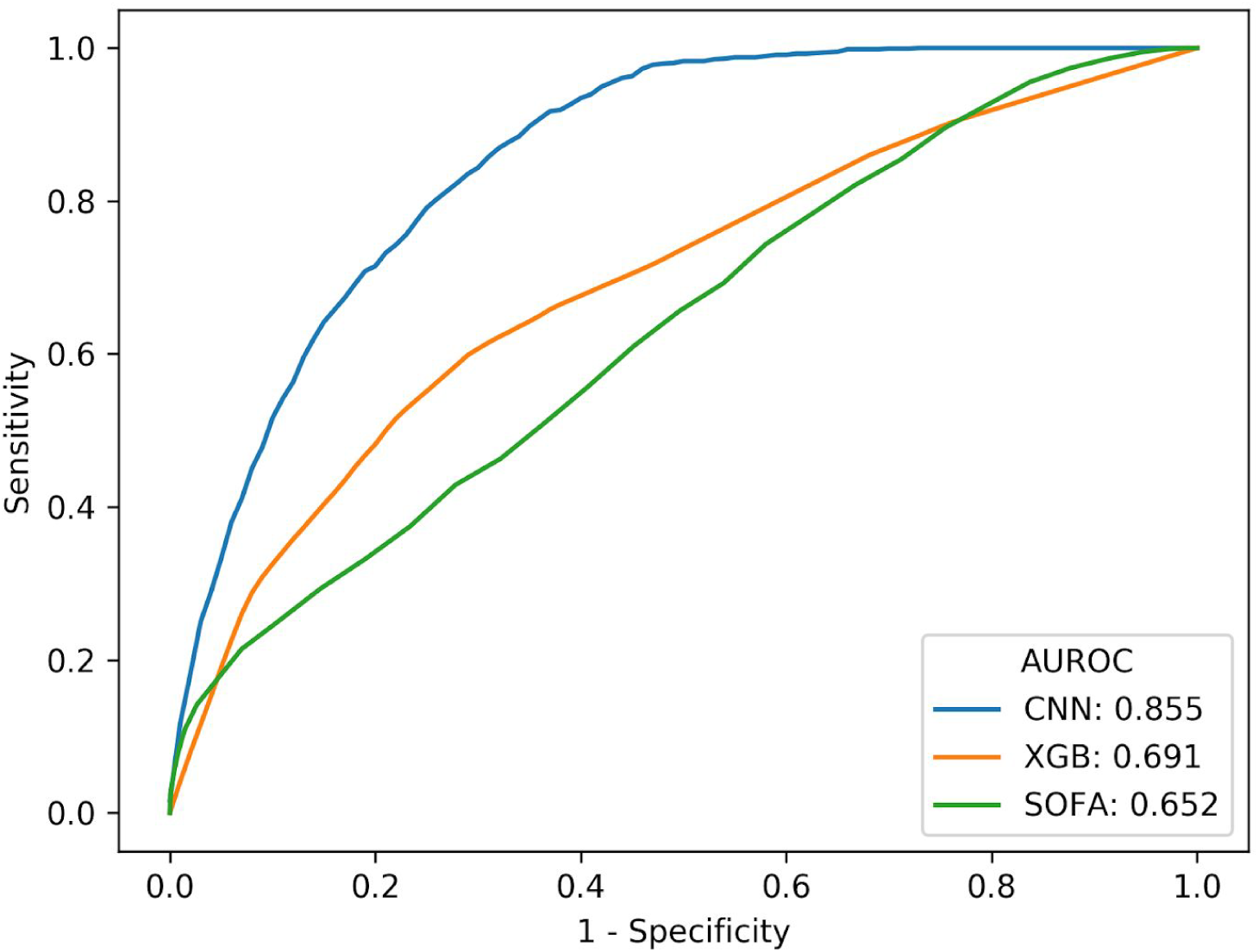
ROC curve comparison of prediction performance using a convolutional neural net (CNN) classifier, an XGBoost (XGB) classifier, and the SOFA score, 72 hours prior to AKI onset on the MIMIC III ICU hold out data set. AUROC, Area Under the Receiver Operating Characteristic curve; SOFA, Sequential Organ Failure Assessment score.

## DISCUSSION

These experiments demonstrate that a convolutional neural network can predict AKI up to 72 hours in advance of KDIGO Stage 2 or Stage 3 AKI onset, with AUROC performance superior to that of an XGBoost classifier and the SOFA scoring system (**Table 2, Figure 2**). Because of the ubiquity of the SOFA score, and previous usage in AKI prediction, it serves as a validated comparator for our current approach [64,65]. The XGBoost comparator is similarly important, primarily due to its broad and successful use in applications to other clinical prediction tasks (e.g., the 2019 Physionet Computing in Cardiology Challenge [66]).

The superiority of the CNN classifier to the XGBoost classifier and the commonly-used SOFA score is evidenced by key performance metrics, such as AUROC and PPV (**Table 2**). The PPV performance improvement is of particular importance. Romero-Brufau *et al*. have argued that AUROC performance may be misleading for clinicians interested in evaluating the clinical impact of a diagnostic tool, as AUROC does not incorporate information about the prevalence of a condition [67]. In fact, for the same reason, AUROC is useful for comparing the performance of tools retrospectively validated on different datasets. This concern regarding PPV and prevalence is relevant to our study, as we found that the prevalence of KDIGO Stages 2 or 3 is roughly 5% in the cohort. The AUROC is a summative metric which may include ranges of operating points which are irrelevant to a given task, whereas PPV can be focused on a clinically relevant operating point. To produce the metrics in **Table 2**, we chose operating points for the CNN and comparators which fixed their sensitivities near 0.80. Zeiberg *et al*. have recently proposed that tools for the prediction of low-prevalence diseases should have PPVs which are at least four times the prevalence, in order to be clinically useful [68]. Of the CNN, XGBoost, and SOFA score, only the CNN would qualify as being clinically useful on the basis of its PPV.

Beyond the text data input through Doc2Vec, CNN predictions were made using only age and 7 routinely collected patient measurements (diastolic blood pressure, systolic blood pressure, temperature, respiratory rate, heart rate, SpO_2_, and Glasgow Coma Scale) as inputs. Importantly, the CNN model did not rely on SCr to make predictions, distinguishing it from other AKI prediction tools. Creatinine levels can take hours or days to rise to AKI thresholds as defined in the KDIGO staging system [69]; therefore, changes in SCr may reflect pre-existing kidney damage. An AKI prediction tool which does not depend on SCr measurements may better afford clinicians the opportunity to intervene early, to prevent AKI development or progression, or to limit further kidney damage. Additionally, using only commonly collected variables in the EHR for AKI prediction allows automatic screening of a general patient population for impending AKI without requiring specialized evaluation.

This study contributes to the growing body of retrospective machine learning literature for the prediction of AKI [70]. Chiofolo et al. (2019) developed a model for AKI prediction and surveillance in ICU patients at a 6-hr prediction window with an AUROC of 0.88 [71]. Fletchet et al. (2017) developed the AKIpredictor, a prognostic calculator for prediction of AKI in ICU patients during the first week of stay [72]. Their KDIGO Stage 2 and 3 model produced AUROCs between 0.77 and 0.84. The AUROC of 0.84 corresponds to a prediction of KDIGO Stage 2 and 3 after gathering 24 hours of data. As a point of comparison, the CNN model used only 5 hours of data before making a prediction. Recent work by Tomasev *et al*. pursued a deep learning approach for continuous risk prediction of deterioration in acute kidney injury patients, and evaluated their tool on a Veteran’s Health Administration dataset of 703,782 adult patients. Algorithm performance at a 48-hour prediction window corresponded to a sensitivity of 55.8% and a specificity of 82.7% [73]. This performance is reported to be “in range” required for regulatory approval [74]. While these studies make important contributions to the domain of AKI research, they depend on the use of SCr to make predictions, which is a lagging marker of kidney function, and most make predictions at shorter prediction horizons than the 72-hour window described in this work.

While the MLA described in this study offers substantial lead time in AKI identification (up to 72 hours), and offers improved predictive performance over our previous work [75], it still requires prospective validation. Additionally, we cannot determine from this retrospective study what impact the algorithm might have on clinicians and their provision of care in clinical settings. Algorithm performance is assessed only on US patients older than age 18, with stays in the ICU, which limits the generalizability of our results to other patient populations and levels of care. Lastly, because there have been several proposed consensus definitions for AKI, the algorithm we described may have different results when compared against non-KDIGO definitions, or in settings which utilize a different standard in their diagnostic procedures.

## CONCLUSION

A convolutional neural network for AKI prediction outperforms XGBoost and the traditional SOFA scoring system, demonstrating superior performance in predicting acute kidney injury up to 72 hours prior to onset, without reliance on measurements of changes in serum creatinine. The use of clinical text data through a Doc2Vec network substantially strengthens prediction performance. Such a tool may improve prediction and early detection of AKI in clinical settings, thereby allowing for earlier intervention.

## Data Availability

The data that support the findings of this study are publicly available from http://www.nature.com/articles/sdata201635.

## Disclosures

### Author’s Contributions

R.D., S.L., and A.A. conceived and designed this study; S.L. and A.A. performed the modeling and statistical analysis; all authors contributed to acquisition, analysis, or interpretation of data; S.L., J.H., A.S., and E.P. drafted the article; all authors revised the article for important intellectual content; and R.D. obtained funding.

## REFERENCES

1. Kashani K, Ronco C. Acute Kidney Injury Electronic Alert for Nephrologist: Reactive versus Proactive? Blood Purif 2016;42:323–328. doi: 10.1159/000450722

2. Al-Jaghbeer M, Dealmeida D, Bilderback A, Ambrosino R, Kellum JA. Clinical Decision Support for In-Hospital AKI. Journal of the American Society of Nephrology. 2017 Nov 2:ASN-2017070765.

3. Hoste EAJ, Bagshaw SM, Bellomo R, Cely CM, Colman R, Cruz DN, Edipidis K, Forni LG, Gomersall CD, Govil D, Honoré PM, Joannes-Boyau O, Joannidis M, Korhonen A-M, Lavrentieva A, Mehta RL, Palevsky P, Roessler E, Ronco C, Uchino S, Vazquez JA, Vidal Andrade E, Webb S, Kellum JA. Epidemiology of acute kidney injury in critically ill patients: The multinational AKI-EPI study. Intensive Care Med 2015; 41: 1411–1423.

4. Wang HE, Muntner P, Chertow GM, Warnock DG: Acute kidney injury and mortality in hospitalized patients. Am J Nephrol 2012;35: 349–355.

5. Uchino S, Bellomo R, Goldsmith D, Bates S, Ronco C: An assessment of the RIFLE criteria for acute renal failure in hospitalized patients. Crit Care Med 2006;34: 1913–1917.

6. Chertow GM, Burdick E, Honour M, Bonventre JV, Bates DW: Acute kidney injury, mortality, length of stay, and costs in hospitalized patients. J Am Soc Nephrol 2005; 16: 3365–3370.

7. Kellum JA, Sileanu FE, Murugan R, Lucko N, Shaw AD, Clermont G: Classifying AKI by urine output versus serum creatinine level. J Am Soc Nephrol 2015; 26: 2231–2238.

8. Hoste EAJ, Kellum JA, Selby NM, Zarbock A, Palevsky PM, Bagshaw SM, Goldstein SL, Cerdá J, Chawla LS. Global epidemiology and outcomes of acute kidney injury. Nat Rev Nephrol. 2018 Oct;14(10):607–625. doi: 10.1038/s41581-018-0052-0. Review. PubMed PMID: 30135570.

9. Freda BJ, Knee AB, Braden GL, Visintainer PF, Thakar CV. Effect of Transient and Sustained Acute Kidney Injury on Readmissions in Acute Decompensated Heart Failure. Am J Cardiol. 2017 Jun 1;119(11):1809–1814.

10. Palevsky PM, Zhang JH, O’Connor TZ, Chertow GM, Crowley ST, Choudhury D, Finkel K, Kellum JA, Paganini E, Schein RMH, Smith MW, Swanson KM, Thompson BT, Vijayan A, Watnick S, Star RA, Peduzzi P. Intensity of renal support critically ill patients with acute kidney injury. New England Journal of Medicine. 2008; 359: 7–20.

11. Kellum JA, Chawla LS, Keener C, Singbartl K, Palevsky PM, Pike FL, Yealy DM, Huang DT, Angus DC. The effects of alternative resuscitation strategies in acute kidney injury patients with septic shock. American Journal of Respiratory and Critical Care Medicine. 2016; 193: 281–287.

12. Khalil P, Murty P, Palevsky PM. The patient with acute kidney injury. Prim Care. 2008 Jun;35(2):239–64, vi. doi: 10.1016/j.pop.2008.01.003. Review. PubMed PMID: 18486715.

13. Forni LG, Dawes T, Sinclair H, et al. Identifying the patient at risk of acute kidney injury a predictive scoring system for the development of acute kidney injury in acute medical patients. Nephron Clinical Practice. 2013;123(3-4): 143–150.

14. Uchino S, Kellum JA, Bellomo R, et al: Beginning and Ending Supportive Therapy for the Kidney (BEST Kidney) Investigators: Acute renal failure in critically ill patients: a multinational, multicenter study. JAMA 2005;294: 813–818.

15. Endre JH, Pickering JW. Acute kidney injury clinical trial design: old problems, new strategies. Pediatr Nephrol. 2013;28: 207–217. doi: 10.1007/s00467-012-2171-3

16. Pickering JW, Ralib AM, Nejat M, Endre ZH. New considerations in the design of clinical trials of acute kidney injury. Clin Invest 2011;1: 637–650.

17. Lachance P, Villeneuve PM, Rewa OG, et al. Association between e-alert implementation for detection of acute kidney injury and outcomes: a systematic review. Nephrol Dial Transplant. 2017;32(2):265–272.

18. Kolhe NV, Staples D, Reilly T et al. Impact of compliance with a care bundle on acute kidney injury outcomes: a prospective observational study. PLoS One 2015; 10: e0132279

19. Terrell KM, Perkins AJ, Hui SL et al. Computerized decision support for medication dosing in renal insufficiency: a randomized, controlled trial. Ann Emerg Med 2010; 56: 623–629

20. Colpaert K, Hoste EA, Steurbaut K et al. Impact of real-time electronic alerting of acute kidney injury on therapeutic intervention and progression of RIFLE class. Crit Care Med 2012; 40: 1164–1170

21. Wilson FP, Shashaty M, Testani J et al. Automated, electronic alerts for acute kidney injury: a single-blind, parallel-group, randomised controlled trial. Lancet 2015; 385: 1966–1974

22. Thomas ME, Sitch A, Baharani J et al. Earlier intervention for acute kidney injury: evaluation of an outreach service and a long-term follow-up. Nephrol Dial Transplant 2015; 30: 239–244

22. Jo S-K, Rosner MH, Okusa MD. Pharmacologic treatment of acute kidney injury: why drugs haven’t worked and what is on the horizon. Clin J Am Soc Nephrol 2007;2: 356–365.

24. Porter CJ, Juurlink I, Bisset LH, Bavakunji R, Mehta RL, Devonald MA. A real-time electronic alert to improve detection of acute kidney injury in a large teaching hospital. Nephrol Dial Transplant. 2014 Oct;29(10):1888–93. doi: 10.1093/ndt/gfu082.

25. Waikar SS, Curhan GC, Wald R et al. Declining mortality in patients with acute renal failure, 1988 to 2002. J Am Soc Nephrol 2006;17: 1143–1150.

26. Xue JL, Daniels F, Star RA et al. Incidence and mortality of acute renal failure in Medicare beneficiaries, 1992 to 2001. J Am Soc Nephrol 2006; 17: 1135–1142

27. Bellomo R, Ronco C, Kellum JA et al. Acute renal failure—definition, outcome measures, animal models, fluid therapy and information technology needs: the Second International Consensus Conference of the Acute Dialysis Quality Initiative (ADQI) Group. Crit Care 2004; 8: R204-R212.

28. Mehta RL, Kellum JA, Shah SV et al. Acute Kidney Injury Network: report of an initiative to improve outcomes in acute kidney injury. Crit Care 2007; 11: R31.

29. Kidney Disease Improving Global Outcomes. Clinical Practice Guideline for Acute Kidney Injury. Kidney Int 2012; 2: 1–138.

30. Palevsky PM, Liu KD, Brophy PD, Chawla LS, Parikh CR, Thakar CV, Tolwani AJ, Waikar SS, Weisbord, SD. KDOQI US Commentary on the 2012 KDIGO Clinical Practice Guideline for Acute Kidney Injury. American Journal of Kidney Diseases. 2013; 61: 649–672.

31. Weisenthal SJ, Quill C, Farooq S, Kautz H, Zand MS. Predicting acute kidney injury at hospital re-entry using high-dimensional electronic health record data. PLoS ONE 2018;13(11): e0204920.

32. Joannidis M, Metnitz B, Bauer P, et al. Acute kidney injury in critically ill patients classified by AKIN versus RIFLE using the SAPS 3 database. Intensive Care Med. 2009; 35: 1692–702.

33. Kolli H, Rajagopalam S, Patel N, et al. Mild acute kidney injury is associated with increased mortality after cardiac surgery in patients with eGFR <60 mL/min/1-73 m(2). Ren Fail. 2010; 32: 1066–72.

34. Wilson FP, Yang W, Feldman HI. Predictors of death and dialysis in severe AKI: the UPHS-AKI cohort. Clin J Am Soc Nephrol. 2013; 8: 527–37.

35. Bihorac A, Delano MJ, Schold JD, et al. Incidence, clinical predictors, genomics, and outcome of acute kidney injury among trauma patients. Ann Surg. 2010; 252: 158–65.

36. Bihorac A, Yavas S, Subbiah S, et al. Long-term risk of mortality and acute kidney injury during hospitalization after major surgery. Ann Surg. 2009; 249: 851–58.

37. Garzotto F, Piccinni P, Cruz D, et al. RIFLE-based data collection/management system applied to a prospective cohort multicenter Italian study on the epidemiology of acute kidney injury in the intensive care unit. Blood Purif. 2011;31:159–71.

38. Newsome BB, Warnock DG, McClellan WM, et al. Long-term risk of mortality and end-stage renal disease among the elderly after small increases in serum creatinine level during hospitalization for acute myocardial infarction. Arch Intern Med. 2008; 168: 609–16.

39. Coca SG, Yusuf B, Shlipak MG et al. Long-term risk of mortality and other adverse outcomes after acute kidney injury: a systematic review and meta-analysis. Am J Kidney Dis 2009; 53: 961–973.

40. Lafrance JP, Miller DR. Acute kidney injury associates with increased long-term mortality. J Am Soc Nephrol 2010; 21: 345–352.

41. Ricci Z, Cruz D, Ronco C. The RIFLE criteria and mortality in acute kidney injury: a systematic review. Kidney Int 2008; 73: 538–546.

42. Uchino S, Kellum JA, Bellomo R et al. Acute renal failure in critically ill patients: a multinational, multicenter study. JAMA 2005; 294: 813–818.

43. Ali T, Khan I, Simpson W et al. Incidence and outcomes in acute kidney injury: a comprehensive population-based study. J Am Soc Nephrol 2007; 18: 1292–1298.

44. Ostermann M, Joannidis M. Acute kidney injury 2016: diagnosis and diagnostic workup. Critical care. 2016 Dec 1;20(1):299.

45. Makris K. The role of the clinical laboratory in the detection and monitoring of acute kidney injury. Journal of Laboratory and Precision Medicine. 2018 Oct 8;3.

46. Davis SE, Lasko TA, Chen G, Siew ED, Matheny ME. Calibration drift in regression and machine learning models for acute kidney injury. J Am Med Inform Assoc. 2017; 24(6):1052–1061.

47. Park S, Ha Baek S, Ahn S, et al. Impact of Electronic Acute Kidney Injury (AKI) Alerts With Automated Nephrologist Consultation on Detection and Severity of AKI: A Quality Improvement Study. Am J Kidney Dis. 2018 Jan; 71(1): 9–19. doi: 10.1053/j.ajkd.2017.06.008

48. de Virgilio C, Kim DY. Transient acute kidney injury in the postoperative period: it is time to pay closer attention. JAMA Surg. 2016;151(5):450–451.

49. Soares DM, Pessanha JF, Sharma A, Brocca A, Ronco C. Delayed nephrology consultation and high mortality on acute kidney injury: a meta-analysis. Blood Purif. 2016;43(1-3):57–67.

50. Thomas ME, Blaine C, Dawnay A, Devonald MA, Ftouh S, Laing C, et al. The definition of acute kidney injury and its use in practice. Kidney Int. 2015;87:62–73.

51. Hodgson LE, Dimitrov BD, Roderick PJ, et al. Predicting AKI in emergency admissions: an external validation study of the acute kidney injury prediction score (APS). BMJ Open 2017;7:e013511.

52. Braitman LE, Davidoff F. Predicting clinical states in individual patients. Ann Intern Med 1996;125:406–12.

53. Feinstein AR. “Clinical Judgment” revisited: the distraction of quantitative models. Ann Intern Med 1994;120:799–805.

54. Concato J, Feinstein AR, Holford TR. The risk of determining risk with multivariable models. Ann Intern Med 1993;118:201–10.

55. Christensen E. Prognostic models including the Child-Pugh, MELD and Mayo risk scores--where are we and where should we go? J Hepatol 2004;41:344–50.

56. Kashani K, Rosner MH, Haase M, Lewington AJP, O’Donoghue DJ, Wilson FP, Nadim MK, Silver SA, Zarbock A, Ostermann M, Mehta RL, Kane-Gill SL, Ding X, Pickkers P, Bihorac A, Siew ED, Barreto EF, Macedo E, Kellum JA, Palevsky PM, Tolwani AJ, Ronco C, Juncos LA, Rewa OG, Bagshaw SM, Mottes TA, Koyner JL, Liu KD, Forni LG, Heung M, Wu VC. Quality Improvement Goals for Acute Kidney Injury. Clin J Am Soc Nephrol. 2019 Jun 7;14(6):941–953. doi: 10.2215/CJN.01250119. Epub 2019 May 17. PubMed PMID: 31101671; PubMed Central PMCID: PMC6556737.

57. Garcia S, Bhatt DL, Gallagher M, Jneid H, Kaufman J, Palevsky PM, Wu H, Weisbord SD. Strategies to Reduce Acute Kidney Injury and Improve Clinical Outcomes Following Percutaneous Coronary Intervention: A Subgroup Analysis of the PRESERVE Trial. JACC Cardiovasc Interv. 2018 Nov 26;11(22):2254–2261. doi: 10.1016/j.jcin.2018.07.044. PubMed PMID: 30466822.

58. MIMIC-III, a freely accessible critical care database. Johnson AEW, Pollard TJ, Shen L, Lehman L, Feng M, Ghassemi M, Moody B, Szolovits P, Celi LA, and Mark RG. Scientific Data (2016). DOI: 10.1038/sdata.2016.35. Available at: http://www.nature.com/articles/sdata201635

59. Desautels T, Calvert J, Hoffman J, Jay M, Kerem Y, Shieh L, Shimabukuro D, Chettipally U, Feldman MD, Barton C, Wales DJ. Prediction of sepsis in the intensive care unit with minimal electronic health record data: a machine learning approach. JMIR medical informatics. 2016;4(3):e28.

60. Le Q, Mikolov T. Distributed representations of sentences and documents. International conference on machine learning 2014 Jan 27 (pp. 1188–1196)

61. Timothy Dozat. Incorporating Nesterov Momentum into Adam. ICLR Workshop, (1):2013–2016, 2016.

62. Chen T, Guestrin C. Xgboost: A scalable tree boosting system. In Proceedings of the 22nd acm sigkdd international conference on knowledge discovery and data mining 2016 Aug 13 (pp. 785–794). ACM.

63. Vincent JL, Moreno R, Takala J, Willatts S, De Mendonça A, Bruining H, Reinhart CK, Suter PM, Thijs LG. The SOFA (Sepsis-related Organ Failure Assessment) score to describe organ dysfunction/failure. On behalf of the Working Group on Sepsis-Related Problems of the European Society of Intensive Care Medicine. Intensive care medicine. 1996 Jul;22(7):707.

64. Hoste EAJ, Clermont G, Kersten A, et al. RIFLE criteria for acute kidney injury is associated with hospital mortality in critically ill patients: a cohort analysis. Crit Care 2006, 10: R73–82. 10.1186/cc4915

65. De Mendonca A, Vincent JL, Suter PM, Moreno R, Dearden NM, Antonelli M, Takala J, Sprung C, Cantraine F. Acute renal failure in the ICU: risk factors and outcomes evaluated by the SOFA score. Intensive Care Med. 2000 July; 26: 915–921.

66. Reyna M, Josef C, Jeter R, Shashikumar S, Westover M, Nemati S, Clifford G, Sharma A. Early prediction of sepsis from clinical data: the PhysioNet/Computing in Cardiology Challenge 2019. Critical Care Medicine. 2019 Oct 14.

67. Romero-Brufau S, Huddleston JM, Escobar GJ, Liebow M. Why the C-statistic is not informative to evaluate early warning scores and what metrics to use. Critical Care. 2015 Dec;19(1):285.

68. Zeiberg D, Prahlad T, Nallamothu BK, Iwashyna TJ, Wiens J, Sjoding MW. Machine learning for patient risk stratification for acute respiratory distress syndrome. PloS one. 2019 Mar 28;14(3):e0214465.

69. Waikar SS, Bonventre JV. Creatinine kinetics and the definition of acute kidney injury. Journal of the American Society of Nephrology. 2009 Mar 1;20(3):672–9.

70. Palevsky PM. Electronic Alerts for Acute Kidney Injury. Am J Kidney Dis. 2018 Jan;71(1):1–2. doi: 10.1053/j.ajkd.2017.09.009. PubMed PMID: 29273153.

71. Chiofolo C, Chbat N, Ghosh E, et al. Automated continuous acute kidney injury prediction and surveillance: a random forest model. Mayo Clin Proc. May 2019;94(5):783–792.

72. Flechet M, Güiza F, Schetz M, et al. AKIpredictor, an online prognostic calculator for acute kidney injury in adult critically ill patients: development, validation and comparison to serum neutrophil gelatinase-associated lipocalin. Intensive Care Med. 2017;43(6):764–773.

73. Tomasev, N. et al. A clinically applicable approach to continuous prediction of future acute kidney injury. Nature 2019;572:116–119.

74. Kellum J, Bihorac A. Artificial intelligence to predict AKI: is it a breakthrough? Nature Reviews Nephrology 2019;15:663–664.

75. Mohamadlou H, Lynn-Palevsky A, Barton C, Chettipally U, Shieh L, Calvert J, Das R. Prediction of Acute Kidney Injury with a Machine Learning Algorithm using Electronic Health Record Data. bioRxiv. 2016 Jan 1: 223354.

